# Accelerated Clonal Hematopoiesis in Pediatric and Young Adult Survivors of Childhood Cancer

**DOI:** 10.1101/2023.05.03.23289475

**Authors:** Chelsee D. Greer, Benjamin R. Kroger, Taylor N. Hartshorne, Jian Xu, Stephen S. Chung, Kathryn E. Dickerson

## Abstract

Clonal hematopoiesis of indeterminate potential(CHIP) is a recognized consequence of aging and a precursor to myelodysplastic syndrome and acute myeloid leukemia which independently increases all-cause mortality in adults. Childhood cancer survivors experience a phenomenon of accelerated aging with increased all-cause mortality; however, the mechanism of this is not known and the prevalence of CHIP not well defined. We prospectively studied 305 pediatric and young adult childhood cancer survivors to determine the prevalence of clonal hematopoiesis(CH). Targeted next-generation sequencing analysis of peripheral blood mononuclear cells elucidated the prevalence of CH (VAF >1%) at a rate of ∼6%, approaching that of adults >50-70 years and much higher than previously reported. This is the first prospective study of CH in pediatric and young adult survivors of childhood cancer and highlights the importance of further investigation to better understand how CH may contribute to treatment-related myeloid neoplasms and other late effects.

## Main

Clonal hematopoiesis(CH) is the process whereby a hematopoietic stem cell acquires a somatic mutation that confers it a growth advantage, thereby producing a clonally distinct subpopulation of hematopoietic cells. With the accumulation of additional mutations, CH can lead hematologic malignancies, such as myelodysplastic syndrome(MDS) and acute myeloid leukemia(AML). Although CH has historically been considered a process tied to aging, it is increasingly recognized to occur in childhood as well when associated with premature aging. With the accumulated progress in curing childhood cancers, there is an expanding cohort of childhood cancer survivors and a critical need to study and ultimately reduce/eliminate the late effects of chemo-radiotherapy. The Childhood Cancer Survivor Study(CCCS) has demonstrated across tumor types a phenomenon of accelerated aging in childhood cancer survivors, culminating in subsequent neoplasms, stroke, and cardiac events—24-year-old survivors of childhood cancer have the same cumulative incidence of grade 3-5 health conditions as 50-year-old siblings.^[1]^ Primary pediatric myeloid neoplasms, including MDS and AML, remain devastating diseases that have a 5-year event free survival(EFS) of ∼40-60%, accounting for nearly 20% of all childhood cancer deaths.^[2-3]^ Additionally, cases of AML and MDS related to prior cancer treatment, referred to as treatment-related myeloid neoplasms(tMN), confer an even poorer prognosis than that of *de novo* AML/MDS, with an overall survival(OS) of 32% and requiring hematopoietic stem cell transplantation(HSCT) for cure.^[4]^ Due to their exposure to chemo-radiotherapy, childhood cancer survivors are at risk of tMN, which predominately occur within two to seven years after their treatment. Currently, tMN is suspected by the presence of a cytopenia(s) with or without leukocytosis on surveillance of peripheral blood counts at scheduled off-therapy visits, making the surveillance for CH easy to implement into off-therapy protocols and may aid in the identification of patients who are at risk of tMN prior to transformation.

Prior whole exome sequencing(WES) studies using a variant allele frequency(VAF) cutoff of 2% demonstrated CH involving presumptive driver genes of hematologic malignancies, such as TET2, DNMT3A, and ASXL1, in 10% of healthy adults >70 years of age and referring to patients meeting these criteria as having clonal hematopoiesis of indeterminate potential (CHIP). ^[5,6,7]^ CHIP is recognized consequence of aging and is well established as a precursor to MDS and AML. ^[5,6,8]^ Adults with CHIP have a 10-fold increased risk of hematologic malignancies and CHIP in adults leads to an increase in all-cause mortality of 40% independent of hematologic malignancies, with the leading cause of death being from atherosclerotic cardiovascular disease and stroke. Furthermore, CHIP likely directly contributes to these phenotypes via elaboration of inflammatory cytokines by macrophages harboring CHIP mutations, as has been demonstrated by studies modeling TET2-mutated CHIP in LDL-receptor atherosclerosis-prone knockout mouse models. ^[5,9]^ Given that subsequent studies have demonstrated significant clinical impacts of clonal hematopoiesis in presumptive drivers of hematologic malignancies at VAFs as low as 1%, particularly in cohorts of adult cancer survivors,^[10]^ we will subsequently refer to this definition of clonal hematopoiesis as CH.

Initial studies evaluating childhood cancer survivors for CH yielded negative results.^[11]^ These efforts had largely been hampered by limited sensitivity of targeted sequencing methodologies, but recent studies using ultra-high-sensitivity error corrected sequencing (ECS) on gDNA from cord blood specimens suggested that CH may occur in children at a much higher rate than could previously be ascertained.^[12]^ Nevertheless, reports of CH in children have been limited to three cases described in non-cancer patients,^[13]^ in children treated for neuroblastoma,^[14]^ in children with solid tumors,^[15]^ and most recently a large cohort of childhood cancer survivors at St Jude Children’s Hospital with a median age of 31.6 years (6.0-66.4) who were a median of 23.5 years from their cancer diagnosis (5.1-55.1 years) and younger than community matched controls that were median of 34.6 years (18.3-72.0 years).^[16]^ All of these previous studies reported an exceedingly low prevalence of CHIP when detection was adjusted to a VAF >2%. Additionally, all studies to date evaluating the presence of pediatric CH in cancer survivors have been retrospective in nature and do not include serial collections following the identification of CH, precluding the ability to examine the clonal dynamics following the emergence of the clone.

Here we report on the first prospective study to determine the prevalence of CH in childhood cancer survivors and demonstrate the feasibility of a novel surveillance strategy for pediatric tMN. This new knowledge may also allow for the identification of patients who have a higher potential to develop other treatment-related toxicities that are associated with premature aging and CHIP, such as cardiovascular events. This approach for surveillance of CH as a precursor to tMN may allow for improvement in the timing of HSCT prior to malignant transformation and ideally inform better prevention strategies that could mitigate the need for HSCT for cure.

We enrolled a representative cohort of pediatric cancer survivors (Fig 1A), and 285 of our 305 consented patients had a CH analysis completed. The age of our patients ranged from three to thirty years (Fig 1B) and time from therapy ranged from two to twenty years (Fig 1C). Patients had a wide variety of chemotherapy exposures (Supp Table 1). We performed targeted sequencing of 24 CH-associated genes on the peripheral blood from the 285 patients and detected 22 mutations in 17 unique patients (Fig 2A). These mutations included nonsynonymous SNV and frameshift mutations in TET2, TP53, DNMT3A, PPM1D, ASXL1, BRCC3, and CBL (Fig 2B) at a composite rate of 5.9% (17 of 285 patients). Out of 62 patients for whom serial specimens were collected, three exhibited growing or diminishing clones over time—a survivor of high-risk(HR) B-cell acute lymphoblastic leukemia(ALL)/lymphoma(Ly), had a diminishing DNMT3A clone detected ∼four years off therapy from 48% to 23% over a one year period during our study, and a survivor of NHL had a diminishing TET2 clone detected ∼four years off therapy from 23% to 15% over a six month period during our study; however, within our dataset a survivor of T-cell ALL/Ly ∼three years off therapy had a growing TET2 clone from <1% to 23% over only a three month period. Only one survivor exhibited multiple CH mutations— a survivor of standard risk(SR) B-cell acute lymphoid leukemia harbored three mutations in CH genes (TET2-2.7%, ASXL1-2%, and TP53-1.7%) detected six years after completion of therapy.

**Figure 1.**
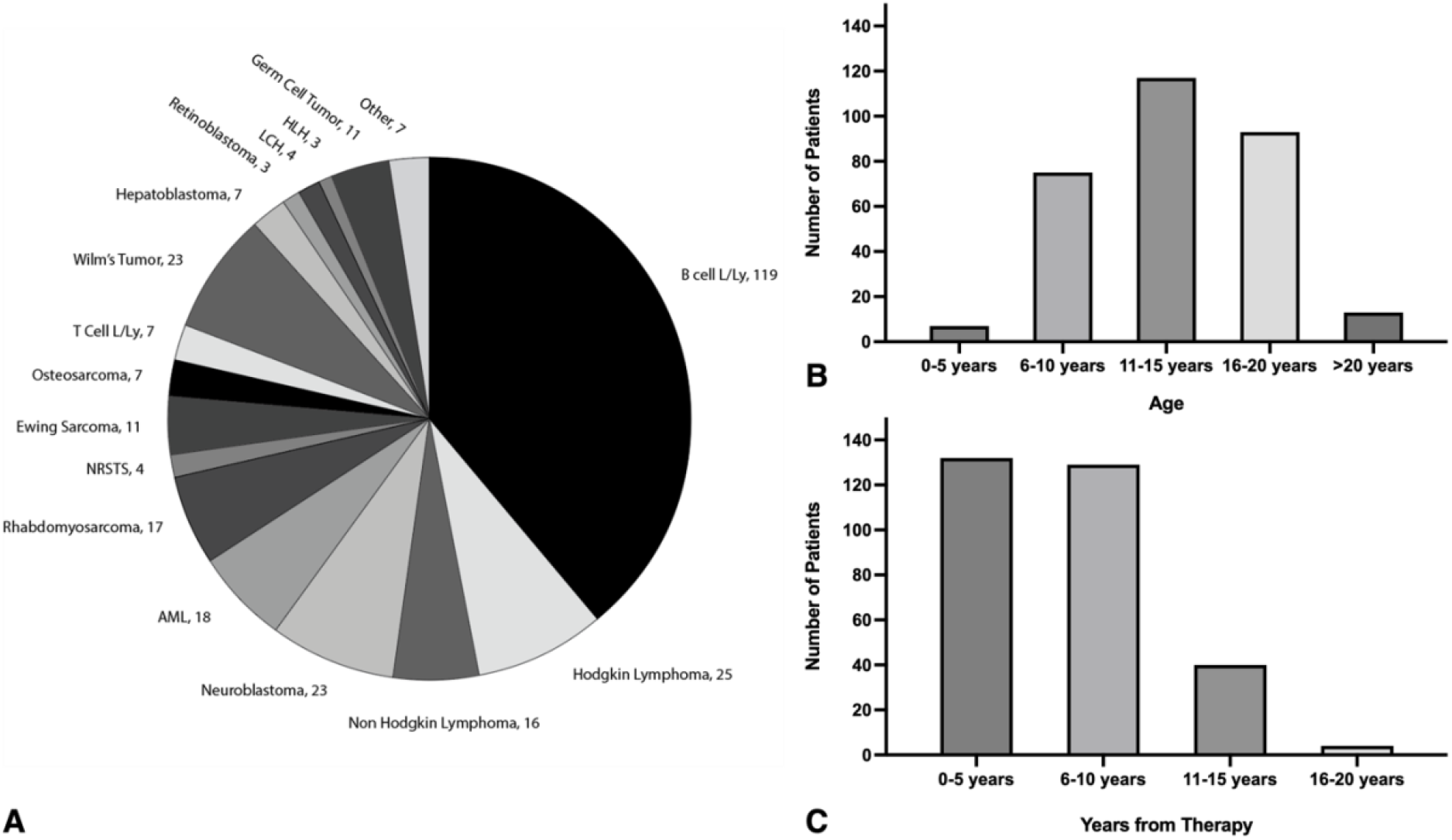
Study population by cancer diagnosis (A), age at diagnosis (B), and years from completion of therapy (C)

**Figure 2.**
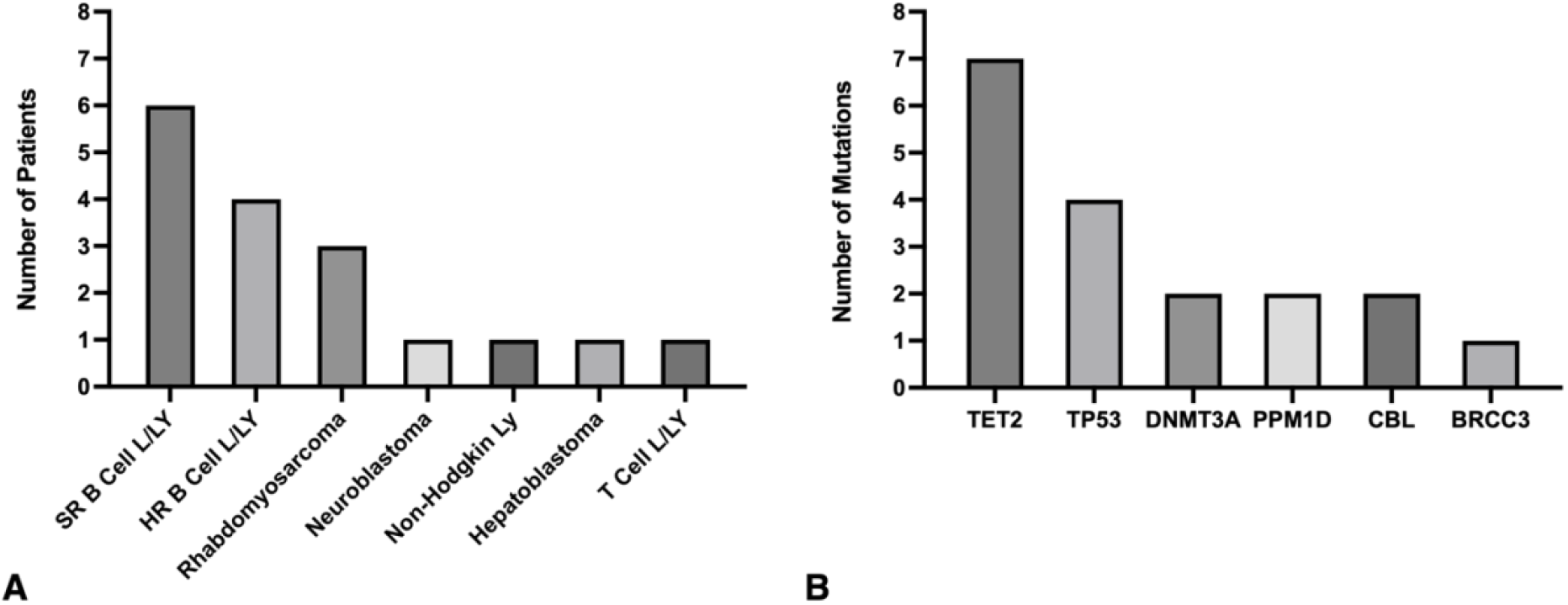
Frequency of CH Mutation in Childhood Cancer Survivors. (A) CH calls by cancer diagnosis. (B) Frequency of CH mutations

TET2 was the most common mutation seen in our cohort as compared to adult studies where DNMT3A mutations predominate. TP53 mutations were the second most common mutation called in our cohort and reflect what has been seen in adult populations exposed to cancer therapy.^[17]^ Given the close proximity in time to treatment (within 1-10 years) of analyzed samples, and the large decrease in the VAF of observed mutations in multiple patients, it seems likely that CH may appear transiently after therapy followed by regression prior to returning with age, with or without progression towards development of a tMN; this is a pattern distinct from what is observed in adults that develop tMN, and it further emphasizes the importance of longitudinal sample collection to better understand the clonal dynamics of CH in childhood cancer survivors. None of our patients harboring CH demonstrate cardiotoxicity or premature ovarian failure, but the detection of CH in childhood cancer survivors across a broad-spectrum of primary diagnoses exemplifies the well-described accelerated aging phenomenon seen in childhood cancer survivors and suggests that ongoing longitudinal studies of may help us to better understand the evolution of risk factors for these negative health effects. Utilizing a paired t-test analysis method, we compared the patients with CH to the patients that did not have CH (Table 1). While no patient characteristics were statistically significant, high rates of CH were noted in rhabdomyosarcoma patients (3 of 14, 21%), there was a slight male predominance in CH, and CH was more likely to be detected in patients closer to their chemotherapy exposures (1-10 years off therapy).

**Table 1.** Characteristics of survivors with CH and without CH. SR = Standard Risk, HR = High Risk, L/Ly = Leukemia/Lymphoma

| Patient Characteristic | CH (n= 17) | No CH (n= 268) | P-value |
| --- | --- | --- | --- |
| **Gender** |  |  |  |
| Male | 10 (58.9%) | 146 (54.5%) | 0.18 |
| Female | 7 (41.1%) | 122 (45.5%) | 0.23 |
| **Ethnicity** |  |  |  |
| White Non-Hispanic | 7 (41.1%) | 97 (36.2%) | 0.28 |
| White Hispanic | 9 (52.9%) | 113 (42.2%) | 0.25 |
| African American | 0 | 29 (10.8%) | 0.42 |
| Asian | 1 (5.8%) | 9 (3.4%) | 0.45 |
| Other/Unknown | 0 | 20 (7.5%) | 0.45 |
| **Age** |  |  |  |
| 1-10 years | 5 (29.4%) | 73 (27.2%) | 0.33 |
| 11-15 years | 8 (47.1 %) | 101 (37.7%) | 0.27 |
| 16-20 years | 2 (11.7%) | 82 (30.6%) | 0.30 |
| Greater than 20 years | 2 (11.7%) | 12 (4.5%) | 0.47 |
| **Diagnosis** |  |  |  |
| SR B Cell L/Ly | 6 (35.3%) | 56 (20.9%) | 0.37 |
| HR B Cell L/Ly | 4 (23.5%) | 44 (16.4%) | 0.39 |
| Rhabdomyosarcoma | 3 (17.6%) | 12 (4.5%) | 0.47 |
| Neuroblastoma | 1 (5.8%) | 21 (7.8%) | 0.44 |
| T Cell L/Ly | 1 (5.8%) | 6 (2.3 %) | 0.48 |
| Non-Hodgkin Ly | 1 (5.8 %) | 15 (5.6%) | 0.46 |
| Hepatoblastoma | 1 (5.8%) | 5 (1.9%) | 0.48 |
| Other Solid Tumors | 0 | 62 (23.1%) | 0.35 |
| Other Liquid Tumors | 0 | 47 (17.5%) | 0.38 |
| **Years Off Therapy** |  |  |  |
| 1-5 years | 9 (52.9%) | 117 (43.7%) | 0.24 |
| 6-10 years | 8 (47.1%) | 110 (41.0%) | 0.25 |
| Greater than 10 years | 0 | 41 (15.3%) | 0.39 |
| **Radiation** |  |  |  |
| Yes | 3 (17.6%) | 66 (24.6%) | 0.34 |
| No | 14 (82.4%) | 220 (82.1%) | 0.06 |
| **BMI** |  |  |  |
| Less than 5th % | 1 (5.8%) | 7 (2.6%) | 0.48 |
| 5th-84th% | 7 (41.2%) | 134 (50.0%) | 0.2 |
| 85- 95th % | 5 (29.4%) | 63 (23.5%) | 0.35 |
| Above 95th% | 4 (23.5%) | 64 (23.9%) | 0.35 |

Although this is a relatively small cohort, and therefore it is prudent to interpret these findings with caution, this is the largest prospective pediatric cohort studied to date. CH is a known consequence of aging, a precursor to myeloid malignancies, and associated with cardiovascular disease. In our prospective, cross-sectional, pilot study of 285 survivors or childhood cancer, we found CH to be present at a rate of 5.9%. This suggests that our patient population has a CH rate over half of that in healthy adults aged 70 years and older. Much less is known about the prevalence in pediatric populations in general; however, the rate of 5.9% is well above existing pediatric studies, which have reported a prevalence ranging from 0.77-1.99%. ^[13,16]^ Furthermore, our study has captured a large Hispanic population that has been under-represented in previous CH studies. Of note, in previous studies CHIP appears to have a lower prevalence in Hispanics but this does not appear to be true in our cohort.^[5]^

In conclusion, childhood cancer survivors demonstrate an increased rate of CH which may be promoted by or potentially a driver of the accelerated aging phenotype observed in this population. Single cell studies will elucidate the cells of interest that harbor CH mutations and if CH mutations are co-occurring within the same cells, while serial sample analysis will define clonal dynamics over time and taken together may inform which clones portend the highest risk of malignant transformation. Given the continued improvement in childhood cancer outcomes and expected growth in this unique population, these results could be of great clinical value. Moreover, if a correlation could be determined between certain treatment-related and/or patient clinical characteristics as risk factors and CH validated as a contributor to cancer late effects such as tMN and others, it may inform a risk-stratification that could be used by clinicians and provide a more robust screening strategy for those defined as higher risk for poor clinical outcome.

## Methods

Patient samples were obtained from participants of the “After the Cancer Experience” (ACE) pediatric survivorship clinic and the adult survivorship clinic via written informed consent on Institutional Review Board-approved research protocols at Children’s Medical Center/UT Southwestern in Dallas in accordance with the Declaration of Helsinki. The ACE survivorship clinic captures ∼85% of the childhood cancer survivors treated at Children’s Medical Center, and patient age, chemotherapy exposure, and late effects are catalogued within an ACE clinic database (Supp Table 1). Eligible patients were identified from the electronic medical record (EMR) at Children’s Medical Center and the UT Southwestern Medical Center. Inclusion criteria included receipt of prior chemo-radiotherapy for a childhood cancer, age less than thirty at time of study enrollment, and being at least one year off-therapy for treatment of cancer. Patients who had undergone allogeneic HSCT and those who had surgery and/or radiation therapy alone were excluded. In total, 305 patients were consented and enrolled from August 2020 to August 2022. Each patient had at least one sample analyzed, and many patients had two to three serial samples collected over the course of the study.

Peripheral blood mononuclear cells (PBMCs) were isolated using a Ficoll-based density centrifugation within 72 hours of collection. Peripheral blood specimens were collected in EDTA coated collection tube and stored at 4°C prior to processing. Genomic DNA (gDNA) was isolated using a Qiagen DNeasy kit, followed by targeted sequencing on a panel of the 24 genes encompassing >95% of previously described recurrent myeloid CHIP mutations (Supp Table 2) through VANTAGE (Vanderbilt University).^[18]^ The mean and median depth of coverage in our study was (698x +/- 177 std dev) and (726x) respectively.

Raw data was aligned to hg38 using Burrow-Wheeler aligner BWA-MEM and processed with the Genome Analysis ToolKit.^[19]^ Single-nucleotide variant calling, and annotation was performed on subsequent BAM files using the CHIP-Detection-Mutect2 Terra Workspace (https://app.terra.bio/#workspaces/terra-outreach/CHIP-Detection-Mutect2). ^[18]^ We selected only those variants which passed Mutect2 filtering and were contained in our whitelist of CH hotspot mutant sites (Supp Table 3) based on established variant calling from the myeloid malignancy field.^[5,14,20]^ Variants were filtered for >50x coverage, had a minimum alternate allele count of 3, variant frequency >1%, and did not occur more than twice in our cohort. Variants with frequency greater than 40% were assumed to be germline unless detected at lower frequency or undetectable in the same patient at a different timepoint.

## Data Availability

All data produced in the present study are available upon reasonable request to the authors

## Supplemental Figures and Figure Legends

**Supp Table 1:**
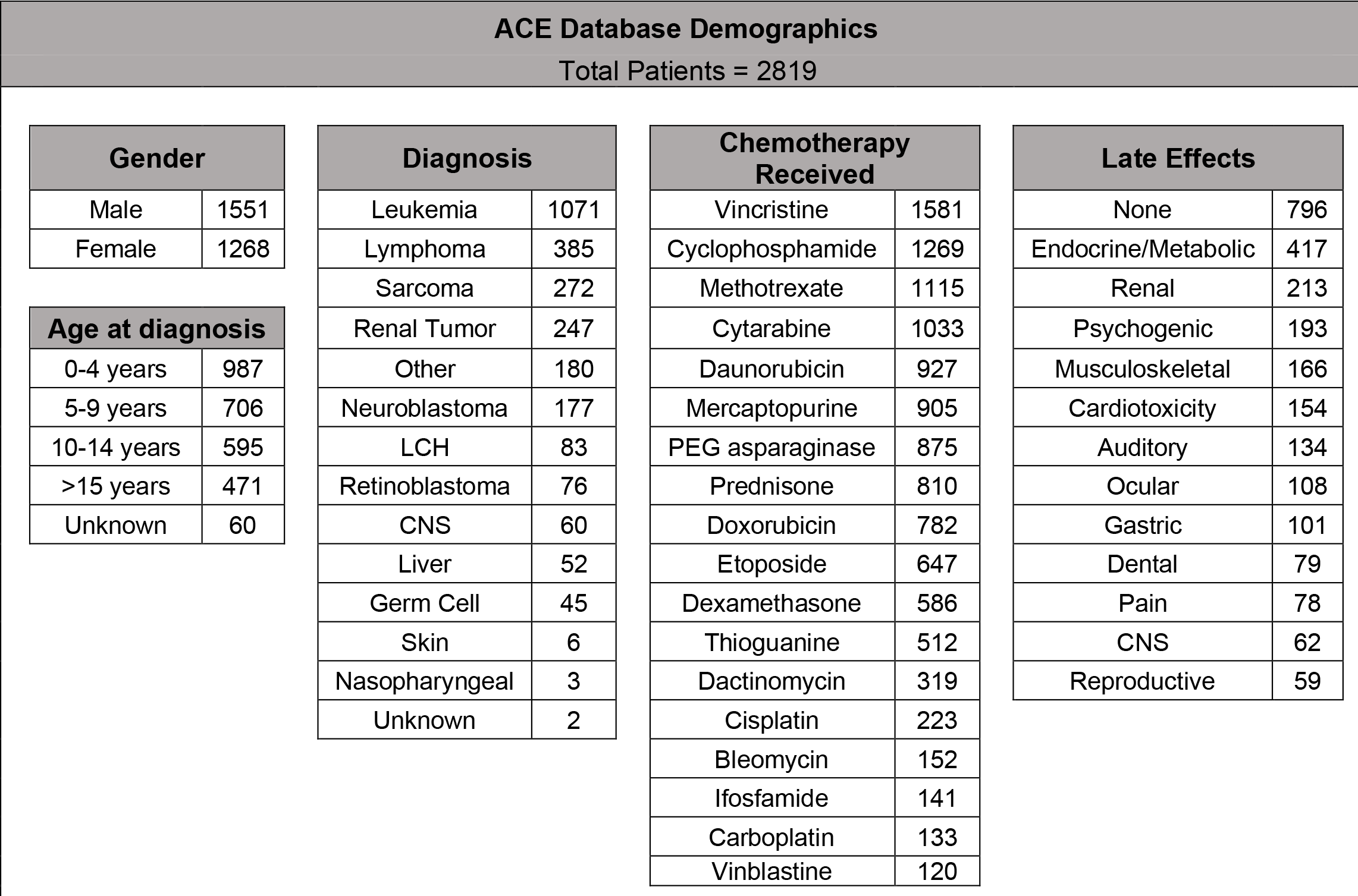
Our Pediatric Cancer Survivorship (“ACE”) Population Demographics at Children’s Health

**Supp Table 2:**
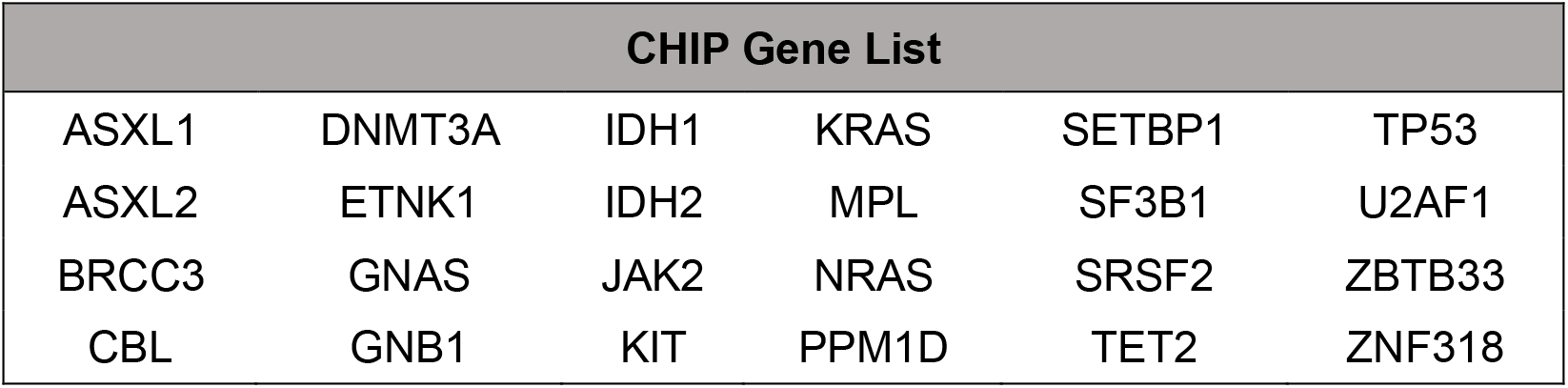
CHIP panel gene list

| CHIP Gene List |  |  |  |  |  |
| --- | --- | --- | --- | --- | --- |
| ASXL1 | DNMT3A | IDH1 | KRAS | SETBP1 | TP53 |
| ASXL2 | ETNK1 | IDH2 | MPL | SF3B1 | U2AF1 |
| BRCC3 | GNAS | JAK2 | NRAS | SRSF2 | ZBTB33 |
| CBL | GNB1 | KIT | PPM1D | TET2 | ZNF318 |

File available for publication

Supp Table 3: CHIP Analysis Whitelist (Adapted from Bick et al,^[18]^ and Feusier et al, Supp Table 4^[13]^)

## References

1. Armstrong, G.T., et al. Aging and risk of severe, disabling, life-threatening, and fatal events in the childhood cancer survivor study. J Clin Oncol, 2014. 32(12): 1218–1227. PMID 24638000

2. Smith, M.A., et al. Outcomes for children and adolescents with cancer: challenges for the twenty-first century. J Clin Oncol, 2010. 28(15): 2625–2634. PMID 20404250

3. Smith, M.A., et al. Declining childhood and adolescent cancer mortality. Cancer, 2014. 120(16): 2497–2506. PMID 24853961

4. Barnard, D.R., et al., Acute myeloid leukemia and myelodysplastic syndrome in children treated for cancer: comparison with primary presentation. Blood, 2002. 100(2): 427–344. PMID 12091332

5. Jaiswal, S. et al. Age-related clonal hematopoiesis associated with adverse outcomes. N Engl J Med 371: 2488–2498 (2014). PMID 2546837

6. Abelson, S. et al. Prediction of acute myeloid leukaemia risk in healthy individuals. Nature 2018 Jul;559(7714):400–404 PMID 29988082

7. Genovese G, et al. Clonal hematopoiesis and blood-cancer risk inferred from blood DNA sequence. N Engl J Med. 2014 Dec 25;371(26):2477–87. PMID 25426838

8. Desai P, et al. Somatic mutations precede acute myeloid leukemia years before diagnosis. Nat Med. 2018 Jul;24(7):1015–1023. PMID 29988143

9. Fuster, J.J., et al. Clonal hematopoiesis associated with TET2 deficiency accelerates atherosclerosis development in mice. Science, 2017. 355(6327): 842–847. PMID 28104796

10. Coombs CC, et al. Therapy-Related Clonal Hematopoiesis in Patients with Non-hematologic Cancers Is Common and Associated with Adverse Clinical Outcomes. Cell Stem Cell. 2017 Sep 7;21(3):374–382. PMID 28803919

11. Collord, G. et al. Clonal haematopoiesis is not prevalent in survivors of childhood cancer. Br J Haematol. 2018 May; 181(4), 537–539. PMID 28369776

12. Wong, W.H., et al. Error-Corrected Sequencing of Cord Bloods Identifies Pediatric AML-Associated Clonal Hematopoiesis. Blood. 2017; 130 (Supplement 1): 2687.

13. Feusier, J.E. et al. Large-scale Identification of Clonal Hematopoiesis and Mutations Recurrent in Blood Cancers. Blood Cancer Discovery. 2021 May;2(3):226–237. PMID 34027416

14. Coorens, T.H. et al. Clonal hematopoiesis and therapy-related myeloid neoplasms following neuroblastoma treatment. Blood. 2021 May 27;137(21):2992–2997. PMID 33598691

15. Spitzer, et al. Bone Marrow Surveillance of Pediatric Cancer Survivors Identifies Clones that Predict Therapy-Related Leukemia. Clin Cancer Res. 2022 April 14;28 (8): 1614–1627. PMID 35078859

16. Hagiwara K, et al. Dynamics of age-versus therapy-related clonal hematopoiesis in long-term survivors of pediatric cancer. Cancer Discov. 2023 Apr 3;13(4):844–857 PMID 36751942

17. Bolton, K, et al. Cancer therapy shapes the fitness landscape of clonal hematopoiesis. Nature Genetics. 2020 Nov;52(11):1219–1226. PMID 33106634

18. Bick AG, et al. Inherited causes of clonal haematopoiesis in 97,691 whole genomes. Nature. 586, 763–768. 2020. PMID 33057201

19. Van der Auwera GA & O’Connor BD. Genomics in the Cloud: Using Docker, GATK, and WDL in Terra. O’Reilly Media (1st Edition; 2020).

20. Jaiswal, S, et al. Clonal Hematopoiesis and Risk of Atherosclerotic Cardiovascular Disease. N Engl J Med. 2017;377(2):111–121. PMID 28636844

